# HIV Prevalence among MSM and TGW in Jamaica Remains High

**DOI:** 10.1101/2025.09.30.25337018

**Authors:** J Peter Figueroa, Carol Jones Cooper, Leshawn Mendoza, Rahanah Khan-Francis, Sharon S Weir

## Abstract

**Introduction:** The study aim was to estimate HIV and syphilis prevalence among men who have sex with a man (MSM) and transgender women (TGW) in Jamaica and assess progress in prevention and treatment.

**Methods:** Participants were recruited sequentially from on-site prevention services, from NGO and government outreach activities, and by peers. Participants were men and TGW aged 16+ who reported sex with a man in the past year and consented to the survey and testing (HIV rapid test, HIV viral load, Bioline syphilis antibody test with TPPA confirmation).

**Results:** Among MSM (n=461), HIV prevalence was 27.1% (95% CI 22.5% - 31.7%), with 81% achieving viral suppression. 29.1% (95% CI 21.9% - 36.2%) were syphilis antibody positive. Within the past 12 months, 22% reported having a sexually transmitted infection (STI) and 60% reported 3+ sex partners. Within the past 6 months 22% received money for sex and 22% were taking pre-exposure prophylaxis (PrEP). 31% used a condom at last sex with their main male partner. 30% were uncomfortable telling anyone they have sex with men. 18% avoided seeking care due to concern they may be treated badly. Factors associated with HIV infection included syphilis (prevalence ratio 3.2 95% CI 2.3-4.6), ever had STI, not completing high school, receiving money for sex, being raped, homeless, jailed/imprisoned. Taking PrEP was protective (prevalence ratio 0.4 95% CI 0.2-0.8).

Among TGW (n=41), HIV prevalence was 53.7% (95% CI 37.3% - 70.0%), with 68% achieving viral suppression. 51.2% (95% CI 37.0% - 65.4%.) were syphilis antibody positive. Within the past 12 months,71% reported 3 or more sex partners. Within the past 6 months 54% received money for sex and 5% were taking PrEP. 32% used a condom at last sex with their main partner. 24% avoided seeking healthcare due to concern they may be treated badly.

**Conclusion:** HIV and syphilis prevalence among MSM and TGW remain high in Jamaica. Adverse life events, syphilis, STI were associated with HIV infection. Increased access to PrEP and syphilis treatment, and structural changes to affirm the rights of MSM and TGW, including changes in social and cultural norms, are needed.

## Introduction

Jamaica has both a generalized and a concentrated HIV epidemic, disproportionately affecting men who have sex with men (MSM) and transgender women (TGW). Adult HIV prevalence in Jamaica was 1.5% in 1996 [1], 1.6% in 2007 [2], 1.8% in 2017 [3] and 1.1% in 2023 [4]. HIV related deaths have declined by 32% since 2010 [4]. An estimated 1,200 adults and children were newly infected with HIV in 2023, a 10% decline since 2010 [4]. Most persons newly HIV infected in 2023 were adult men, many of whom were MSM [4].

Historically, HIV prevalence in Jamaica has been high among MSM: in 1986 9.6% [6], by 1995 32.3%, in 2007 32.9% [7], in 2011 31.4% [8], and in 2017 32.9% [9] (Table 1). The 876 study in 2017 was the first to make a distinction between HIV prevalence in MSM (29.6%) and TGW (51.0%) [9]. In Jamaica, same-sex intimacy, transgender and gender diverse people are stigmatized, and the Offences Against the Person Act criminalizes anal sex, further perpetuating discrimination against MSM and TGW. Despite numerous educational campaigns homosexuality and HIV remain highly stigmatized although somewhat less than previously [5-9]. These findings highlight the ongoing challenges in addressing HIV in Jamaica.

**Table 1.** HIV Prevalence among MSM in Jamaica.

| Year | No. of MSM surveyed | HIV prevalence | Reference |
| --- | --- | --- | --- |
| 1986 | 125 | 9.6% | Murphy, Gibbs, Figueroa et al. JAIDS 1988 [6] |
| 1995 | 201 | 32.3% | Rossi unpublished |
| 2007 | 207 | 32.9% | Figueroa, Weir, Jones-Cooper et al. WIMJ 2012 [7] |
| 2011 | 449 | 31.4% | Figueroa, Jones Cooper, Edwards et al. PLOS 2015 [8] |
| 2017 | 652 MSM, TGW, TGM<br>537 MSM,<br>102 TGW, 13 TGM | 32.9% (MSM & TGW)<br>29.6% (MSM)<br>51.0% (TGW) | 876 study MOHW report 2018 unpublished [9] |

The National Program needs current information to improve programs and reduce HIV infection among MSM and TGW. The aim of this study was to estimate the prevalence of HIV and other sexually transmitted infections (STI), associated risk factors and behaviours.

## Methods

Participants were recruited sequentially at locations in Jamaica providing targeted prevention services to MSM or TGW from 16 July 2024 to 2 November 2024. Treatment sites were excluded and refusals monitored. Respondents had to be born male, 18 years or older, or 16-17 years and not with parents or on a family errand and had anal sex with a man in the past year. Transgender women were defined as a person born with male biologic sex who identifies as female gender. Persons who described themselves as “transgender” were included as TGW even if they also self-identified as a man.

Participants gave informed written consent and agreed to answering a questionnaire and being tested for HIV and syphilis, including viral load and TRUST antigen if positive for HIV and reactive for syphilis, respectively. Unique participant identification numbers linked questionnaire, biological specimens, and laboratory results. Names and contact information were requested and stored separately in a master log accessible only to the core study team and was only used for providing laboratory results and referring to care. Participants who completed the survey were given a US$6.50 phone card and US$16 cash as a contribution towards transportation and their time. Participants were offered 3-5 appointment cards and invitation letters to invite their peers and the study coordinator’s contact information. Participants were not given any financial incentive for recruiting a peer. The Ministry of Health and Wellness (MOHW) Ethics Committee approved the study.

The Principal Investigator (JPF) and Study Coordinator (CJC) were well known to NGO and MOHW staff providing services and to the MSM and TGW communities. The Study Coordinator supervised a trained team of experienced women interviewers. A structured questionnaire designed and uploaded on tablets to a secure server was administered face-to-face. Questions asked about socio-demographic status, HIV awareness, risk perception, sexual behaviour, condom use, STI symptoms, health seeking behaviour, drug use and social vulnerability. Participants had the option to self-complete especially sensitive questions. Interviews took place privately at NGOs, MOHW workshops, health centres and mutually agreed sites.

HIV testing was done using the Determine rapid test. If a participant tested positive on Determine test a dried blood sample was taken on plasma separation cards for viral load testing using Roche PCR. Syphilis testing was done using Bioline with confirmation of positive results using Treponema pallidum particle agglutination assay (TPPA). A positive syphilis test was interpreted as “ever infected”. Participants with a positive TPPA and a TRUST titre of 8 or higher were considered currently infected. Confidential results were given to participants free of cost, with counselling and referral to treatment as indicated.

Socio-economic status (SES) of participants was assessed by the interviewer on a scale of 1-10 with 1-3 being lowest SES and 7-10 highest.

All study protocols maintained confidentiality and privacy to avoid unwanted exposure of participants.

## Data analysis

The study supervisors and coordinator reviewed all questionnaires completed by interviewers for quality control prior to submission to the server. Interviewers were consulted to clarify any answers that appeared inconsistent. 29 clusters of respondents were identified based on the organisation facilitating recruitment, approach taken (facility or outreach/workshop), location (parish) and month recruited. Participants presenting appointment cards were assigned to the same cluster as the participant who told them about the survey.

Although none of the sociodemographic or behavioral variables except schooling were associated with age (when categorized in 3 groups 16-24, 25-34 and 35+ years), because age was so strongly associated with prevalence HIV infection, we included age in generalized linear models to estimate prevalence ratios and 95% Confidence Intervals (CI). We used proc genmod (SAS Institute, Cary NC), controlled for age and recruitment location (outreach or facility), assumed a binomial distribution, used the link=log option, and treated the cluster as a repeated subject.

## Results

### MSM

Four (4) persons refused to do the survey, six (6) turned away before being approached, 17 were ineligible, 64 had already participated including 24 who were deliberately attempting to participate in the survey twice. Of 461 MSM study participants, 193 (41.9%) were recruited through services at NGO facilities and 268 (58.1%) through outreach/workshop activities of NGOs or parish health staff (Figure 1). 53 were recruited by peers already interviewed. 48% were aged 20-29 years while 12% were 16-19 years (Table 2). Most (75%) were assessed by interviewers to be truthful in their answers. 59% were recruited from Kingston & St Andrew or St James parish (11%).

**Table 2.**
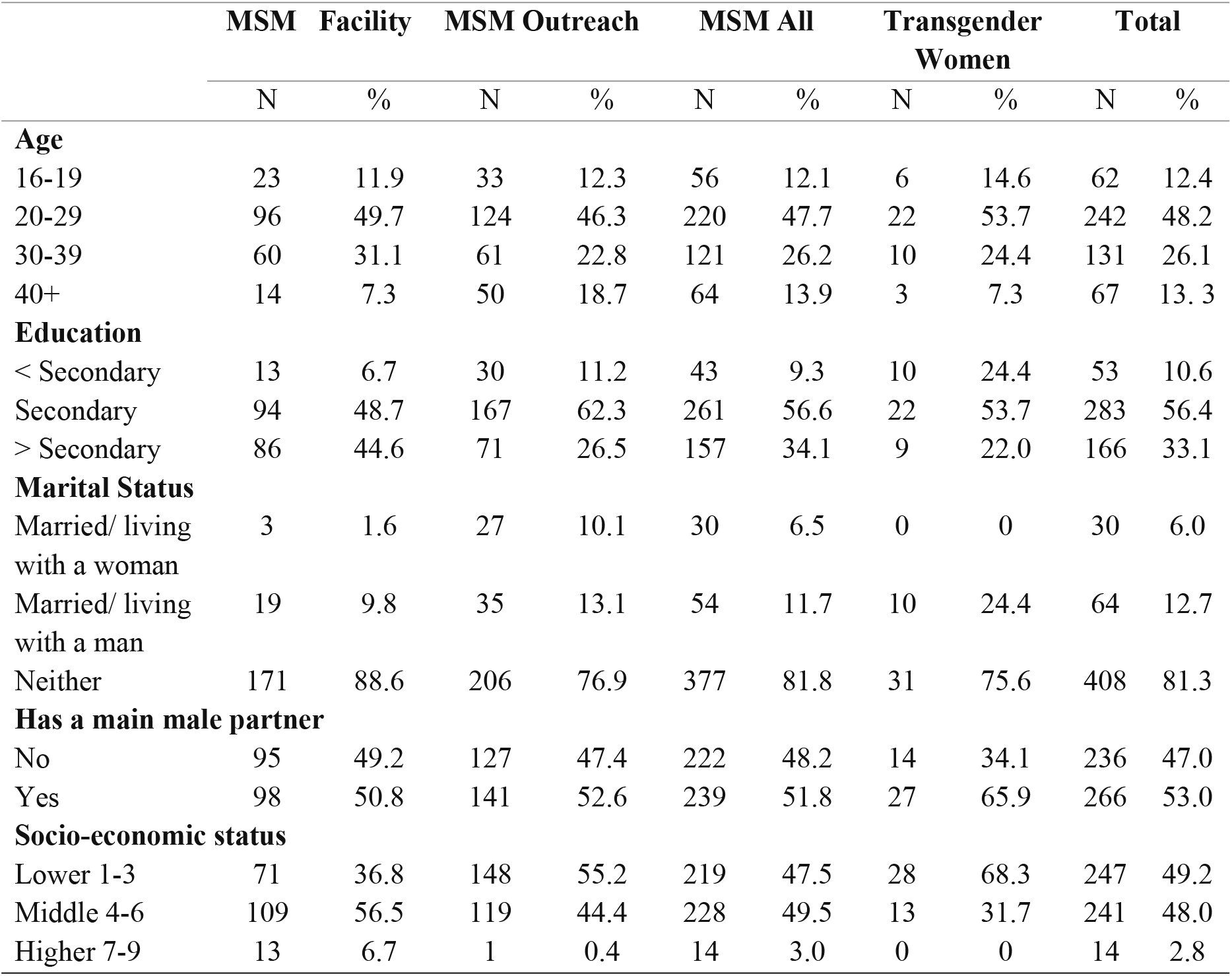
Sociodemographic Characteristics of MSM and TGW in Jamaica 2024.

**Figure 1.**
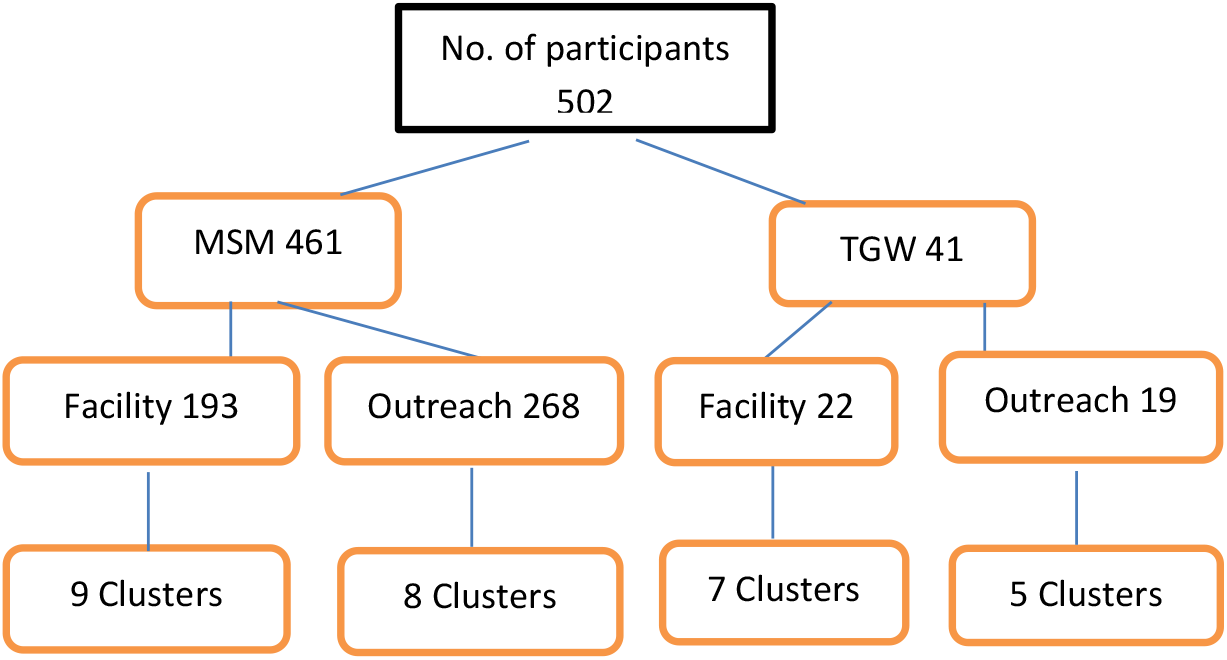
Recruitment Flow Diagram using BBS-Lite Approach.

Although 90.7% of MSM had completed secondary school including 34.1% who had completed tertiary education, 47.5% were assessed as very low SES and only 3% as high SES.

A third of MSM (30.4%) had sex before age 16. Fewer than 20% were married or living with a partner (male or female). MSM recruited through outreach were more likely to report sex with both men and women than those recruited through facilities (42.9% vs. 23.3%).

HIV prevalence was 27.1% (95% CI 22.5% - 31.7%), increasing from 10.7% among teens to 38.9% among MSM 30 years or older (Figure 2).

**Figure 2.**
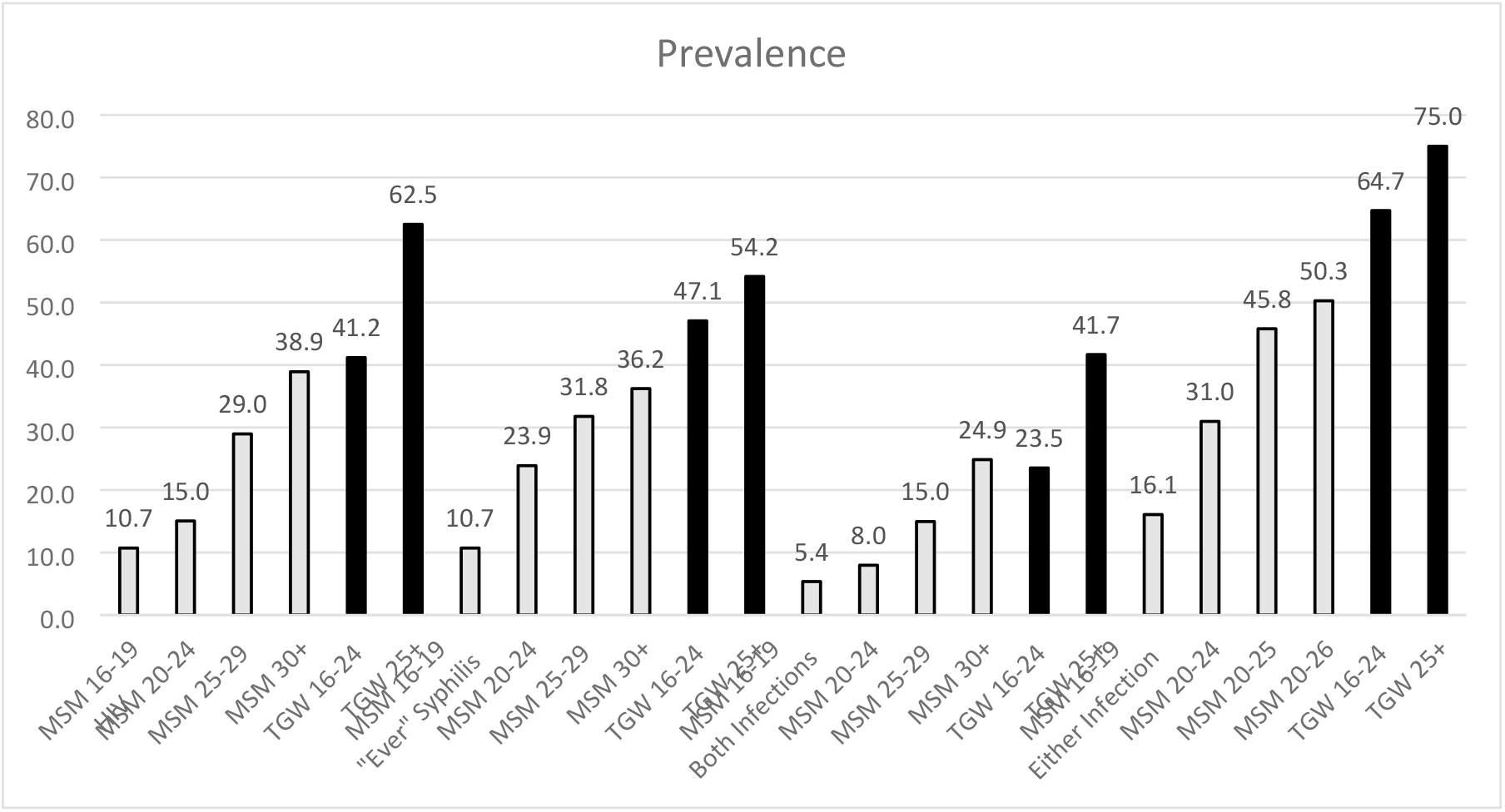
Prevalence of HIV and Syphilis (past and current) among MSM in Jamaica 2024.

HIV prevalence was significantly higher among those recruited through outreach than facility (31.7%, vs. 20.7%) (Table 3). Among those with HIV infection, 74% reported their HIV+ status to the interviewer and 70% reported taking anti-retroviral treatment (ART). Under the assumption that those with viral load suppression were on ART and knew their status, we estimate that 91% knew their status, 89% were on treatment and 81% were virally suppressed. Among HIV-ve participants, most (65%) assessed that they had no or little risk of becoming HIV infected.

**Table 3.** Prevalence of HIV and Syphilis and HIV Treatment Cascade among MSM & TGW in Jamaica 2024.

|  | MSM Facility |  |  |  | MSM Outreach |  |  |  | Transgender Women |  |  |  |
| --- | --- | --- | --- | --- | --- | --- | --- | --- | --- | --- | --- | --- |
|  | N | % | CI |  | N | % | CI |  | N | % | CI |  |
| **HIV Test** |  |  |  |  |  |  |  |  |  |  |  |  |
| Positive | 40 | 20.7 | 16.2 | 25.2 | 85 | 31.7 | 25.3 | 38.1 | 22 | 53.7 | 37.3 | 70.0 |
| Negative | 153 | 79.3 | 74.8 | 83.8 | 183 | 68.3 | 61.9 | 74.7 | 19 | 46.3 | 30.0 | 62.7 |
| Total | 193 | 100.0 |  |  | 268 | 100.0 |  |  | 41 | 100.0 |  |  |
| **Among HIV+** |  |  |  |  |  |  |  |  |  |  |  |  |
| Knows status | 38 | 95.0 | 84.7 | 100.0 | 76 | 89.4 | 81.9 | 96.9 | 19 | 86.4 | 67.8 | 100.0 |
| On ART | 37 | 92.5 | 83.0 | 100.0 | 74 | 87.1 | 79.8 | 94.3 | 19 | 86.4 | 67.8 | 100.0 |
| Suppressed | 34 | 85.0 | 69.1 | 100.0 | 67 | 78.8 | 67.7 | 87.9 | 15 | 68.2 | 39.1 | 97.3 |
| Total* | 40 | 100.0 |  |  | 85 | 100.0 |  |  | 22 | 100.0 |  |  |
| **Syphilis Test** |  |  |  |  |  |  |  |  |  |  |  |  |
| Reactive | 35 | 18.1 | 8.4 | 27.8 | 99 | 36.9 | 28.1 | 45.7 | 21 | 51.2 | 37.0 | 65.4 |
| Non-reactive | 158 | 81.9 | 72.2 | 91.6 | 169 | 63.1 | 54.3 | 71.9 | 20 | 48.8 | 34.6 | 63.0 |
| Total | 193 | 100.0 |  |  | 268 | 100.0 |  |  | 41 | 100.0 |  |  |
| **Among Reactive** |  |  |  |  |  |  |  |  |  |  |  |  |
| Acute | 9 | 25.7 | 6.5 | 45.0 | 30 | 30.3 | 20.2 | 40.4 | 8 | 38.1 | 14.9 | 61.2 |
| Non-acute | 26 | 74.3 | 55.0 | 93.5 | 69 | 69.7 | 59.6 | 79.8 | 13 | 61.9 | 38.8 | 85.1 |
| Total | 35 | 100.0 |  |  | 99 | 100.0 |  |  | 21 | 100.0 |  |  |

\*1 excluded due to missing viral load.

Syphilis prevalence (current or past infection, n=134) was 29.1% (95% CI 21.9% - 36.2%), increasing from 10.7% among teens to 36.2% among MSM aged 30 and older; and significantly higher among those recruited by outreach (37.0% vs 18.1%). 8.5% (39) of MSM had current syphilis.

Most MSM (81.3%) reported visiting a public health clinic. Approximately half reported a health worker had told them they had a STI including 21.9% who were told in the past 12 months. When asked about symptoms within the past 3 months, 7.6% reported a genital discharge, 4.6% a genital ulcer, 2% genital warts and 9.5% burning on urination. Of those with symptoms, 39.0% reported sex while symptomatic.

MSM met sex partners online (87.8%); at parties (56.8%); on the street (51.9%); at bars and clubs (39.2%). Approximately one third (36.4%) had accepted money for sex including 22.1% in the past 6 months (Table 4). More admitted to accepting money for sex (24% vs 21%) when answering the question privately on the tablet than face-to-face with the interviewer. 16.5% of MSM reported having five or more one-night stands in the past 12 months. 31.9% reported two or more sex partners in the past 4 weeks.59.9% reported three (3) or more male sex partners in the past 12 months. 29.1% of MSM believed that their main sex partner had sex with another person. 38.4% had never had sex with a woman. Among those who had, 66.9% used a condom the last time they had sex with a woman. 51.2% of MSM said their partner used a condom the last time they had receptive anal intercourse. 24.1% of MSM said that they were never bottom. 25.6% of MSM said that they had ever taken pre-exposure prophylaxis (PrEP) (Table 5). 87% had heard about PrEP and 41% were interested in learning more. HIV prevalence was lower among those who had ever taken PrEP (12% vs 32.3%). 100 MSM (21.7%) said that they took PrEP in the past 6 months, 63 (13.7%) took it continuously and 37 (8%) had stopped.

**Table 4.** Sexual Behavior of MSM and TGW in Jamaica 2024.

|  | MSM Facility |  | MSM Outreach |  | MSM All |  | TGW |  | Total |  |
| --- | --- | --- | --- | --- | --- | --- | --- | --- | --- | --- |
|  | N | % | N | % | N | % | N | % | N | % |
| **Age at first sex** |  |  |  |  |  |  |  |  |  |  |
| < Age 16 | 50 | 25.9 | 90 | 33.6 | 140 | 30.4 | 18 | 43.9 | 158 | 31.5 |
| Age 16+ | 143 | 74.1 | 178 | 66.4 | 321 | 69.6 | 23 | 56.1 | 344 | 68.5 |
| **Sex with only men or both men and women** |  |  |  |  |  |  |  |  |  |  |
| Only men | 148 | 76.7 | 153 | 57.1 | 301 | 65.3 | 38 | 92.7 | 339 | 67.5 |
| Both men & women | 45 | 23.3 | 115 | 42.9 | 160 | 34.7 | 3 | 7.3 | 163 | 32.5 |
| **Male partners past 4 weeks** |  |  |  |  |  |  |  |  |  |  |
| 0 | 41 | 21.2 | 56 | 20.9 | 97 | 21.0 | 4 | 9.8 | 101 | 20.1 |
| 1 | 86 | 44.6 | 131 | 48.9 | 217 | 47.1 | 13 | 31.7 | 230 | 45.8 |
| 2-4 | 53 | 27.5 | 64 | 23.9 | 117 | 25.4 | 14 | 34.1 | 131 | 26.1 |
| 5+ | 13 | 6.7 | 17 | 6.3 | 30 | 6.5 | 10 | 24.4 | 40 | 8.0 |
| **New partners past 4 weeks** |  |  |  |  |  |  |  |  |  |  |
| 0 | 118 | 61.1 | 185 | 69.0 | 303 | 65.7 | 13 | 31.7 | 316 | 62.9 |
| 1 | 46 | 23.8 | 54 | 20.1 | 100 | 21.7 | 17 | 41.5 | 117 | 23.3 |
| 2+ | 29 | 15.0 | 29 | 10.8 | 58 | 12.6 | 11 | 26.8 | 69 | 13.7 |
| **Male partners past 12 months** |  |  |  |  |  |  |  |  |  |  |
| 1 | 37 | 19.2 | 72 | 26.9 | 109 | 23.6 | 6 | 14.6 | 115 | 22.9 |
| 2 | 36 | 18.7 | 39 | 14.6 | 75 | 16.3 | 6 | 14.6 | 81 | 16.1 |
| 3-5 | 60 | 31.1 | 86 | 32.1 | 146 | 31.7 | 13 | 31.7 | 159 | 31.7 |
| 6-10 | 35 | 18.1 | 41 | 15.3 | 76 | 16.5 | 4 | 9.8 | 80 | 15.9 |
| 11+ | 25 | 13.0 | 29 | 10.8 | 54 | 11.7 | 12 | 29.3 | 66 | 13.1 |
| **Number 1 night stands past 12 months** |  |  |  |  |  |  |  |  |  |  |
| 0 | 68 | 35.2 | 104 | 38.8 | 172 | 37.3 | 15 | 36.6 | 187 | 37.3 |
| 1 | 30 | 15.5 | 55 | 20.5 | 85 | 18.4 | 7 | 17.1 | 92 | 18.3 |
| 2-4 | 56 | 29.0 | 72 | 26.9 | 128 | 27.8 | 6 | 14.6 | 134 | 26.7 |
| 5+ | 39 | 20.2 | 37 | 13.8 | 76 | 16.5 | 13 | 31.7 | 89 | 17.7 |
| **Believes main male partner had sex with another person past 6 months** |  |  |  |  |  |  |  |  |  |  |
| Yes | 57 | 29.5 | 77 | 28.7 | 134 | 29.1 | 14 | 34.1 | 148 | 29.5 |
| No | 35 | 18.1 | 49 | 18.3 | 84 | 18.2 | 7 | 17.1 | 91 | 18.1 |
| Don't know | 6 | 3.1 | 15 | 5.6 | 21 | 4.6 | 6 | 14.6 | 27 | 5.4 |
| No main male partner | 95 | 49.2 | 127 | 47.4 | 222 | 48.2 | 14 | 34.1 | 236 | 47.0 |
| **Received cash for sex** |  |  |  |  |  |  |  |  |  |  |
| Never | 135 | 69.9 | 158 | 59.0 | 293 | 63.6 | 14 | 34.1 | 307 | 61.2 |
| Not in the past 6 months | 24 | 12.4 | 42 | 15.7 | 66 | 14.3 | 5 | 12.2 | 71 | 14.1 |
| Within the past 6 months | 34 | 17.6 | 68 | 25.4 | 102 | 22.1 | 22 | 53.7 | 124 | 24.7 |

**Table 5.** Risk Assessment, Health Care Seeking and Prevention among MSM and TGW in Jamaica 2024.

|  | MSM Facility |  | MSM Outreach |  | MSM All |  | TGW |  | Total |  |
| --- | --- | --- | --- | --- | --- | --- | --- | --- | --- | --- |
|  | N | % | N | % | N | % | N | % | N | % |
| **To what extent are you at risk of HIV infection** |  |  |  |  |  |  |  |  |  |  |
| No chance | 26 | 13.5 | 40 | 14.9 | 66 | 14.3 | 6 | 14.6 | 72 | 14.3 |
| Little chance | 92 | 47.7 | 87 | 32.5 | 179 | 38.8 | 5 | 12.2 | 184 | 36.7 |
| Moderate chance | 32 | 16.6 | 47 | 17.5 | 79 | 17.1 | 9 | 22.0 | 88 | 17.5 |
| Good chance | 15 | 7.8 | 34 | 12.7 | 49 | 10.6 | 8 | 19.5 | 57 | 11.4 |
| Already HIV+ | 28 | 14.5 | 60 | 22.4 | 88 | 19.1 | 13 | 31.7 | 101 | 20.1 |
| **Used condom last time with main male partner** |  |  |  |  |  |  |  |  |  |  |
| Yes | 52 | 26.9 | 89 | 33.2 | 141 | 30.6 | 13 | 31.7 | 154 | 30.7 |
| No | 46 | 23.8 | 52 | 19.4 | 98 | 21.3 | 14 | 34.1 | 112 | 22.3 |
| No main male partner | 95 | 49.2 | 127 | 47.4 | 222 | 48.2 | 14 | 34.1 | 236 | 47.0 |
| **Partner used condom last receptive anal sex** |  |  |  |  |  |  |  |  |  |  |
| Yes | 89 | 46.1 | 147 | 54.9 | 236 | 51.2 | 26 | 63.4 | 262 | 52.2 |
| No | 58 | 30.1 | 56 | 20.9 | 114 | 24.7 | 15 | 36.6 | 129 | 25.7 |
| Never receptive | 46 | 23.8 | 65 | 24.3 | 111 | 24.1 | 0 | 0 | 111 | 22.1 |
| **Knowledge & Use of PrEP** |  |  |  |  |  |  |  |  |  |  |
| Never heard of PrEP | 8 | 4.1 | 52 | 19.4 | 60 | 13.0 | 3 | 7.3 | 63 | 12.5 |
| Heard of PrEP, not interested | 33 | 17.1 | 60 | 22.4 | 93 | 20.2 | 13 | 31.7 | 106 | 21.1 |
| Heard of and interested | 70 | 36.3 | 120 | 44.8 | 190 | 41.2 | 19 | 46.3 | 209 | 41.6 |
| Used over 6 months ago | 10 | 5.2 | 8 | 3.0 | 18 | 3.9 | 1 | 2.4 | 19 | 3.8 |
| Used during past 6 months but not continuously | 25 | 13.0 | 12 | 4.5 | 37 | 8.0 | 3 | 7.3 | 40 | 8.0 |
| Continuous use 6 months | 47 | 24.4 | 16 | 6.0 | 63 | 13.7 | 2 | 4.9 | 65 | 12.9 |
| **Ever been to a public health clinic** |  |  |  |  |  |  |  |  |  |  |
| Yes | 144 | 74.6 | 231 | 86.2 | 375 | 81.3 | 36 | 87.8 | 411 | 81.9 |
| No | 49 | 25.4 | 37 | 13.8 | 86 | 18.7 | 5 | 12.2 | 91 | 18.1 |
| **Told by a health worker in past 12 months that you have an STI?** |  |  |  |  |  |  |  |  |  |  |
| Yes | 39 | 20.2 | 62 | 23.1 | 101 | 21.9 | 9 | 22.0 | 110 | 21.9 |
| No | 153 | 79.3 | 206 | 76.9 | 359 | 77.9 | 32 | 78.0 | 391 | 77.9 |

Where percentage distributions do not sum to 100, there are 1-2 cases of missing data.

Many MSM were abused or suffered as children: 54.5% were beaten; 47.9% were verbally abused or neglected; 28.4% were sexually abused; and 43.5% witnessed their mother or someone else verbally or physically abused. As adults, 28.8% had slept outdoors or in a shelter because they were homeless. 25.8% had been raped and 27.7% had been violently attacked by a sexual partner. Approximately 11% said that their partner had occasionally or often raped or forced them to have sex (9.8% and 1.1%) (Table 6). HIV prevalence was higher among MSM who ever reported being raped than those who were never raped (38.1% vs 23.3%).

**Table 6.** Adverse Events and Stigma among MSM and TGW in Jamaica 2024.

|  | MSM Facility |  | MSM Outreach |  | MSM All |  | Transgender Women |  | Total |  |
| --- | --- | --- | --- | --- | --- | --- | --- | --- | --- | --- |
|  | N | % | N | % | N | % | N | % | N | % |
| **How big a problem is:** |  |  |  |  |  |  |  |  |  |  |
| ***Stigma from health care workers*** |  |  |  |  |  |  |  |  |  |  |
| Big Problem | 18 | 9.3 | 26 | 9.7 | 44 | 9.5 | 7 | 17.1 | 51 | 10.2 |
| Small Problem | 23 | 11.9 | 23 | 8.6 | 46 | 10.0 | 9 | 22.0 | 55 | 11.0 |
| Not A Problem | 152 | 78.8 | 219 | 81.7 | 371 | 80.5 | 25 | 61.0 | 396 | 78.9 |
| ***Being rejected by family*** |  |  |  |  |  |  |  |  |  |  |
| Big Problem | 55 | 28.5 | 84 | 31.3 | 139 | 30.2 | 27 | 65.9 | 166 | 33.1 |
| Small Problem | 45 | 23.3 | 55 | 20.5 | 100 | 21.7 | 7 | 17.1 | 107 | 21.3 |
| Not A Problem | 92 | 47.7 | 129 | 48.1 | 221 | 47.9 | 7 | 17.1 | 228 | 45.4 |
| ***Being abused because of your sexual orientation*** |  |  |  |  |  |  |  |  |  |  |
| Big Problem | 60 | 31.1 | 85 | 31.7 | 145 | 31.5 | 29 | 70.7 | 174 | 34.7 |
| Small Problem | 38 | 19.7 | 46 | 17.2 | 84 | 18.2 | 5 | 12.2 | 89 | 17.7 |
| Not A Problem | 95 | 49.2 | 137 | 51.1 | 232 | 50.3 | 7 | 17.1 | 239 | 47.6 |
| ***Being physically attacked because of your sexual orientation*** |  |  |  |  |  |  |  |  |  |  |
| Big Problem | 60 | 31.1 | 80 | 29.9 | 140 | 30.4 | 28 | 68.3 | 168 | 33.5 |
| Small Problem | 38 | 19.7 | 42 | 15.7 | 80 | 17.4 | 5 | 12.2 | 85 | 16.9 |
| Not A Problem | 95 | 49.2 | 146 | 54.5 | 241 | 52.3 | 8 | 19.5 | 249 | 49.6 |
| ***How frequently does your partner insult, harass, embarrass or try to control you?*** |  |  |  |  |  |  |  |  |  |  |
| Never | 125 | 64.8 | 172 | 64.2 | 297 | 64.4 | 19 | 46.3 | 316 | 62.9 |
| Occasionally | 35 | 18.1 | 50 | 18.7 | 85 | 18.4 | 6 | 14.6 | 91 | 18.1 |
| Often | 13 | 6.7 | 19 | 7.1 | 32 | 6.9 | 12 | 29.3 | 44 | 8.8 |
| No Partner | 20 | 10.4 | 27 | 10.1 | 47 | 10.2 | 4 | 9.8 | 51 | 10.2 |
| ***How frequently does your partner slap hit or choke you?*** |  |  |  |  |  |  |  |  |  |  |
| Never | 151 | 78.2 | 201 | 75.0 | 352 | 76.4 | 23 | 56.1 | 375 | 74.7 |
| Occasionally | 15 | 7.8 | 34 | 12.7 | 49 | 10.6 | 10 | 24.4 | 59 | 11.8 |
| Often | 7 | 3.6 | 6 | 2.2 | 13 | 2.8 | 4 | 9.8 | 17 | 3.4 |
| No Partner | 20 | 10.4 | 27 | 10.1 | 47 | 10.2 | 4 | 9.8 | 51 | 10.2 |
| ***How frequently has your partner raped or forced you to have sex?*** |  |  |  |  |  |  |  |  |  |  |
| Never | 155 | 80.3 | 208 | 77.6 | 363 | 78.7 | 31 | 75.6 | 394 | 78.5 |
| Occasionally | 15 | 7.8 | 30 | 11.2 | 45 | 9.8 | 4 | 9.8 | 49 | 9.8 |
| Often | 2 | 1.0 | 3 | 1.1 | 5 | 1.1 | 2 | 4.9 | 7 | 1.4 |
| No Partner | 21 | 10.9 | 27 | 10.1 | 48 | 10.4 | 4 | 9.8 | 52 | 10.4 |

227 Where percentage distributions do not sum to 100, there are 1-2 cases of missing data.

Most MSM (71%) said that they drank alcohol less than weekly or never, 21% said they drank weekly, and 8% said they drink daily. 33% said they had consumed six (6) or more alcoholic drinks on an occasion in the past 4 weeks. Marijuana use was common. In the past 6 months no participant had used crack/ cocaine or injected drugs, 5% reported ecstasy use and 37% used some other drug.

Nearly all (94%) said it was easy to access an HIV test and 95% said that they had ever been tested (Table 7). 75% had been tested within the past 12 months. Of these, 97% received their result, and 77% were counselled. Many shared their HIV result with their main partner (52%), or a friend or co-worker (51%). 71.6% had heard of the HIV self-testing kit and 24% had ever used it.

**Table 7.** HIV Testing by HIV Status among MSM and TGW in Jamaica 2024.

|  | MSM Facility |  | MSM Outreach |  | MSM All |  | TGW |  | Total |  |
| --- | --- | --- | --- | --- | --- | --- | --- | --- | --- | --- |
|  | N | % | N | % | N | % | N | % | N | % |
| **HIV Testing** |  |  |  |  |  |  |  |  |  |  |
| Reports test is easy to get | 183 | 94.8 | 251 | 93.7 | 434 | 94.1 | 39 | 95.1 | 473 | 94.2 |
| Tested in past 12 months | 29 | 15.0 | 63 | 23.5 | 92 | 20.0 | 17 | 41.5 | 109 | 21.7 |
| Ever used self-test kit | 51 | 26.4 | 59 | 22.0 | 110 | 23.9 | 12 | 29.3 | 122 | 24.3 |
| **HIV Testing by HIV Status** | 142 | 73.6 | 209 | 78.0 | 351 | 76.1 | 29 | 70.7 | 380 | 75.7 |
| *Among HIV Negative:* |  |  |  |  |  |  |  |  |  |  |
| No test in past 12 months | 24 | 15.7 | 50 | 27.3 | 74 | 22.0 | 5 | 26.3 | 79 | 22.3 |
| Tested in the past 12 months | 129 | 84.3 | 133 | 72.7 | 262 | 78.0 | 14 | 73.7 | 276 | 77.7 |
| Total | 153 | 100.0 | 183 | 100.0 | 336 | 100.0 | 19 | 100.0 | 355 | 100.0 |
| *Among HIV Positive:* |  |  |  |  |  |  |  |  |  |  |
| Knows HIV status | 38 | 95.0 | 76 | 89.4 | 114 | 91.2 | 19 | 86.4 | 133 | 89.9 |
| Does not know status and no test in past 12 months | 1 | 2.5 | 3 | 3.5 | 4 | 3.2 | 2 | 9.1 | 6 | 4.1 |
| Does not know status and tested in past 12 months | 1 | 2.5 | 6 | 7.1 | 7 | 5.6 | 1 | 4.5 | 8 | 5.4 |
| Total | 40 | 100.0 | 85 | 100.0 | 125 | 100.0 | 22 | 100.0 | 147 | 100.0 |

Most MSM (59%) were able to answer all five (5) knowledge questions about HIV correctly. Nearly all participants knew that a healthy-looking person can be infected with HIV and knew that using condoms reduces risk of HIV transmission. There was no clear association between knowledge of HIV and HIV prevalence although prevalence was higher among persons thinking that condoms do not reduce the risk of HIV transmission (40.7% vs 26.2%).

One third of MSM (30.1%) were not comfortable telling anyone that they have sex with men and 34.5% were comfortable telling less than half the persons they knew. 9.1% of MSM said that in the past year they had been treated badly by a health worker in a public clinic while 17.6% of MSM had avoided seeking care because they were worried that they may be treated badly. Being rejected by their family was considered a big problem (30%), as was being abused (31%) or attacked (30%) because of their sexual orientation.

Always using PrEP in the past 6 months (14%) was protective against HIV for MSM (prevalence ratio 0.3 95% CI 0.1-0.6) (Table 8). Completing high school or tertiary education, and aged 16 years or older at first sex were suggestive but not statistically significant.

**Table 8.** Prevalence Ratios for association with HIV among MSM in Jamaica 2024.

|  | **Yes**<br>**% HIV+** | **No**<br>**% HIV+** | **Prevalence**<br>**Ratio\*** | **95% CI** | **P value** |
| --- | --- | --- | --- | --- | --- |
| Age at first sex >16 years | 23.8 | 35.0 | 0.7 | 0.5– 1.1 | 0.11 |
| Completed high school or more | 24.6 | 51.2 | 0.7 | 0.4 – 1.2 | 0.18 |
| Always used PrEP past 6 months | 7.9 | 30.2 | 0.3 | 0.1– 0.6 | <0.01 |
| Ever used PrEP | 11.9 | 32.4 | 0.4 | 0.2 – 0.8 | <.01 |
| Ever had Syphilis | 55.2 | 15.6 | 3.2 | 2.3 – 4.6 | < 0.0001 |
| Ever told had STI | 42.6 | 22.8 | 1.8 | 1.4 – 2.4 | < 0.0001 |
| 3+ one-night stands past year | 33.1 | 24.4 | 1.4 | 1.1 – 1.9 | <0.05 |
| 11+ sex partners past year | 30.2 | 26.8 | 1.1 | 0.7 – 1.5 | 0.79 |
| Ever raped | 38.1 | 23.3 | 1.8 | 1.3 – 2.5 | <0.001 |
| Slept outdoors because homeless | 37.9 | 22.8 | 1.6 | 1.2 – 2.1 | <0.001 |
| Ever violently attacked by sex partner | 36.8 | 24.1 | 1.5 | 1.2 – 1.9 | <0.0001 |
| Ever received money for sex | 38.0 | 21.1 | 1.8 | 1.3 – 2.6 | <0.001 |
| Received money for sex in past 6 months | 37.6 | 24.7 | 1.5 | 1.1 – 2.1 | <01 |

\* Prevalence ratios and CI are estimates from models that stratified by recruitment cluster and controlled for age, except for the variable “ever violently attacked by a sex partner” where the model including age did not converge.

Factors significantly associated with HIV infection in MSM were ever having syphilis (prevalence ratio 3.2 95% CI 2.3-4.6), being told by a health provider that they ever had STI or had STI in the past 12 months, ever raped, ever spent a night in jail or prison, ever homeless, ever receiving money for sex or receiving money for sex in the past six (6) months.

### TGW

41 TGW were recruited, 19 though outreach and 22 from a facility. Three (3) were recruited by peers. Half (53.7%) were aged 20-29 years while 15% were 16-19 years. Most (18) were recruited from Kingston and St Andrew, but at least one was recruited from all but four parishes.

87% self-identified themselves. Three (3) TGW self-identified as men but described themselves as transgender and were counted as TGW. Most (75.6%) were neither married nor living with a partner. Compared with MSM, fewer TGW reported sex with women (7.3% vs 34.6%), fewer TGW completed secondary school (75.7% vs 90.7%), and more were in the lowest SES 1-3 category (68.3% vs 47.5%).

Among TGW, HIV prevalence was 53.7% (95% CI 37.7%-69.6%); current or past syphilis infection was 51.2% (95% CI 37.0%-65.4%) and current syphilis was 38.1%, all significantly higher than MSM. Among 22 TGW with HIV infection, 14 (64%) reported their HIV+ status to the interviewer and 14 (64%) reported taking ART. Under the assumption that those with viral load suppression were on ART and knew their status, we estimate that 86% knew their status, 86% were on treatment and 68% were virally suppressed. Among HIV-ve TGW, 10 of 19 (53%) assessed that they had no or little risk of becoming HIV infected.

56.1% of TGW had ever been told they had a STI, including 22% who had been told within the past 12 months. Most (87.8%) said that they had ever attended a public health clinic.

TGW reported meeting partners online (97.6%), at parties (78%), on the street (73%), and at bars and clubs (52.2%). 61% had ever accepted money for sex and 54% did so in the past 6 months. 71% reported having three (3) or more male partners in the past 12 months. 58.6% reported two (2) or more male sex partners in the past 4 weeks. Significantly more TGW than MSM had a new male sex partner in the past 4 weeks (68.3% vs 34.2%). 63.4% of TGW said that their most recent male partner used a condom.

Although almost all had heard about PrEP, only 6 TGW (14.6%) had ever taken PrEP. HIV prevalence was lower among those who had ever taken PrEP (16.7% vs 60.0%).

Prevalence of adverse events was higher among TGW: 68.3% were beaten as children; 63.4% were verbally abused or neglected; 46.3% were sexually abused; and 55.0% witnessed their mother or someone else verbally or physically abused. As adults, 73.2% had slept outdoors or in a shelter; 53.7% had been raped and 61.0% had been violently attacked by a sexual partner. TGW reported alcohol and drug use similar to MSM.

Among HIV-ve TGW, 73.7% had been tested within the past 12 months. Most TGW (75.6%) had heard of the HIV self-testing kit and 38.7% had ever used it. Knowledge of HIV was similar to that of MSM. 26.8% of TGW reported being treated badly by a health worker in a public clinic in the past year and 24.4% had avoided seeking care because they were worried that they may be treated badly. Perceived big problems included family rejection (66%) and abuse (71%) or being attached because of sexual orientation (68%).

## Discussion

HIV prevalence among MSM and TGW remains high in Jamaica in 2024 (MSM 27.1% 95% CI 22.5% - 31.7%, TGW 53.7% 95% CI 37.7% - 69.6%). HIV and syphilis prevalence among MSM increases rapidly by age. The HIV prevalence remains similarly high for the past 3 decades. The factors contributing to this include the social vulnerability of MSM in Jamaica due to many adverse life events, widespread stigma and discrimination including the Offenses against the Person Act which criminalizes anal sex, risk behaviour including unprotected sex with multiple sex partners, sex in exchange for money, other STI particularly syphilis, and low perception of risk.

As many as 50% of participants reported that a health provider had ever told them that they had a STI and 22% were so informed in the past 12 months. HIV prevalence was high among persons reporting ever having a STI (MSM 45.0%, TGW 69.6%) or having a STI in the past 12 months (MSM 42.6%, TGW 66.7%). Many participants tested positive for ever having syphilis (MSM 29.1%, TGW 51.2%) and syphilis was significantly associated with HIV (prevalence ratio among MSM 3.2 95% CI 2.3-4.6). Ready access to prompt diagnosis and treatment of syphilis and other STIs is essential. Syphilis testing should be routinely offered to all MSM and young adults at emergency departments of hospitals, health centres and NGOs providing services to MSM and TGW.

Risk behaviours among MSM and TGW continue to be common: most participants had multiple sex partners; 37.6% said that they had ever received cash for sex and 23.6% had done so in the past 6 months. Among the latter, HIV prevalence was high (MSM 38%, TGW 55%).

Many participants suffered adverse life events as children and/or adults. HIV prevalence was higher among MSM who had been raped (38%) or homeless (38%). Experiencing adverse life events was associated with increased HIV prevalence in the 2011 survey in Jamaica (8) and it appears to be the case in this survey also.

Stigma against MSM and TGW remains an important problem in Jamaica. TGW were more likely than MSM to be verbally abused, beaten or sexually abused as a child and raped, homeless, or ever attacked by a sex partner. Parents must be educated to recognize that beating and abusing children affects them negatively while praising good behaviour and expressing love for your child promotes self-esteem and appropriate behaviour [10]. There must be safe places established in schools and communities for MSM and TGW, and counselors trained to offer them support and guidance. Most MSM (81%) and TGW (88%) had attended a public health clinic. However, many (MSM 11.2%, TGW 30.6%) said that they had been treated badly by a health care worker in the past year and many (MSM 17.7%, TGW 14.4%) said they had avoided seeking health care at a public clinic because they were worried that they would be treated badly.

Despite the continued high HIV prevalence there are two encouraging developments. HIV prevalence among MSM aged 16 – 24 years declined from 20.7% in 2011 [8] to 18% in 2017 [9] and 12.5% in 2024. Also, HIV prevalence among MSM using pre-exposure prophylaxis (PrEP) was 12% compared with 32% among those not using PrEP (prevalence ratio 0.4 95% CI 0.2-0.8). The reduction in social interaction associated with the COVID-19 pandemic may have contributed to lower HIV prevalence among young MSM in 2024 or the sample may have been biased towards including those on PrEP. It is also possible that in this cross-sectional study the lower prevalence of HIV among PrEP users reflects the PrEP requirement of being HIV negative rather than a decreased HIV incidence among PrEP users. Nevertheless, a clear priority must be to get many more MSM and TGW using PrEP.

HIV prevalence among MSM participants was significantly higher among MSM recruited at outreach activities than those recruited at NGO facilities. This may be because MSM recruited through outreach were more likely to report receiving cash for sex (40.2% vs 29.0%) and those recruited at service sites were more likely to be using PrEP (42.5% vs 13.2%). The socio-economic status, knowledge of HIV, number of sexual partners or adverse life events were similar between the two recruitment groups. 502 participants were recruited within four (4) months, which was swifter than previous surveys. However, the study team assessed that many MSM and TGW were not being reached, and too few workshops were being conducted.

It is difficult to assess how representative a sample of the MSM and TGW populations was achieved. Many TGW were willing to be interviewed but avoided recruitment because of the blood test. Limitations of the survey include the inability to recruit a representative sample and the small number of TGW and participants of higher social status enrolled. Strengths of the study include swift completion, comprehensive data collected, high quality control, analysis of data adjusted by strata and clusters, and recruitment consistent with the UNAIDS/WHO BBS-Lite method [11] which recruits through NGOs and public health staff providing HIV prevention and other services.

## Conclusion

While there are encouraging signs such as a reduction in HIV prevalence among MSM 16 – 24 years and the introduction of PrEP, we are failing to achieve a significant reduction in HIV prevalence among MSM and TGW. PrEP use must be expanded and supported by innovative programs to improve HIV prevention, treatment and psycho-social care. It is disheartening to witness the unacceptable high level of verbal, physical and sexual abuse that MSM and TGW receive as children and the high level of rape, homelessness and violence they suffer as adults. Structural changes are needed to reduce stigma and discrimination and affirm the rights of MSM and TGW, including amendment of the Offences Against the Person Act and changes in social and cultural norms.

## Authors’ contributions

JPF, SSW and CJC designed the study; JPF, CJC, RFK and LM implemented the study; SSW, JPF and CJC analyzed the data; JPF drafted the manuscript; all authors edited the manuscript and accepted the final submission.

## Acknowledgements

The study team thanks the Ministry of Health and Wellness, A-Z Information Jamaica Ltd, NGOs and The Global Fund.

## Data Availability Statement

The Ministry of Health and Wellness, Kingston, Jamaica houses the database devoid of any personal identifiers.

